# Evaluation of the Ultima Genomics UG 100 sequencer for low-cost, high-sensitivity metagenomic pathogen detection from cerebrospinal fluid

**DOI:** 10.1101/2025.05.16.25327810

**Authors:** Ryan C. Shean, Keith D. Tardif, Alexandra Rangel, Jacob Dutrschi, David Bogumil, Alix Cruse, Samm Hernadez, Nika Iremadze, Sarah Pollock, Doron Lipson, Benjamin T. Bradley

## Abstract

Metagenomic next-generation sequencing (mNGS) is a diagnostic tool allowing near universal pathogen detection directly from clinical specimens. Despite promising clinical data, broad adoption of mNGS has been hindered by high cost and reduced sensitivity relative to targeted nucleic acid amplification tests (NAATs). Recently, Ultima Genomics revealed the UG 100 NGS platform which advertises 10 billion reads per $2,400 sequencing wafer. By significantly lowering costs and improving sequencing depth, the historical value proposition of mNGS may be improved. This study evaluates the UG 100 sequencer for metagenomic pathogen detection from cerebrospinal fluid in suspected cases of meningitis and encephalitis. Of 28 specimens with a pathogen identified by routine clinical testing, reads matching to the known pathogen were identified in 93% (26/28) of cases. Near full-length genomes were recovered for three organisms (human herpesvirus-1, *Streptococcus pneumonia*, and *Haemophilus influenzae*), with the ability to detect putative antimicrobial resistance genes for *H. influenzae*. Recovery of *Borrelia burgdorferi* reads (6.1 RPM and 9.03 RPM) was achieved from clinical samples with late cycle threshold values (39.7 and 43.0, respectively). Limit of detection (LOD) studies demonstrated detection of HSV-1 and *S. pneumoniae* at 50 and 5 copies/mL, respectively, which is below the reported limit of detection for the orthogonal NAATs used in this study. Reducing sequencing costs and improving the analytical sensitivity removes two major hurdles for mNGS adoption by clinical laboratories. While these results are preliminary, they demonstrate a future in which mNGS may be more widely implemented.

**Importance:** Metagenomic next-generation sequencing has struggled to gain wider adoption for nearly a decade, due in part to concerns related to its high cost and reduced performance versus targeted molecular assays. This study demonstrates the ability of the UG100 sequencing platform to significantly reduce metagenomic sequencing costs (to approximately $12 per 50M reads) while maintaining highly sensitive pathogen detection rates. Improvements to cost and analytical performance may shift clinical metagenomics from an expensive test-of-last-resort to a front-line diagnostic for identifying infections.

## Introduction

Metagenomic next-generation sequencing (mNGS) is a diagnostic methodology that attempts to sequence millions of nucleic acid fragments in parallel from clinical samples to detect pathogens^1^. This technique has been shown to have several advantages over traditional microbiologic methods. One such advantage is the ability to provide “hypothesis free” detection for a range of pathogens (bacteria, viruses, fungi, protozoa, and eukaryotic parasites) without explicit inclusion in the ordering physician’s differential^2^.

While initially met with enthusiasm, the broad adoption of mNGS has been hindered by technical and financial limitations. One of the most well-studied applications, CSF mNGS has shown a clinical impact in 28-57% of suspected meningitis and encephalitis (M/E) cases^3–7^. This variability stems partly from differences in how clinical significance is defined as well as from the reduced sensitivity of current mNGS methods as compared to targeted molecular assays like real-time PCR^3^.

Financially, mNGS adoption is limited by the high testing cost—approximately $2,000 per test—and the restricted availability of the assay, which is offered mainly through select reference laboratories and companies. To address these issues, many laboratories have implemented stewardship strategies to prioritize testing for individuals with the highest pre-test probability of actionable results^8^. For mNGS to achieve wider adoption, testing costs must be reduced and assay sensitivity must be improved.

Recently, a novel sequencing platform has been developed that significantly reduces the per base sequencing cost. Utilizing a proprietary reagent delivery platform to massively scale reaction chemistry, the Ultima Genomics UG 100 sequencer (Ultima Genomics, Fremont CA, USA) provides ∼10 billion reads per run at a sequencing wafer cost of ∼$2400^9,10^. This technology has previously been evaluated for high throughput applications in human genomics such as single cell RNA-Seq^11^, whole transcriptome sequencing^12^, and cell-free circulating tumor detection^13^. Reducing the sequencing cost of mNGS has the potential to increase adoption of clinical metagenomics. For example, if the cost of metagenomic testing could approach that of syndromic testing by multiplex PCR panels, the historical use of mNGS as a “test of last resort” may shift to earlier in the diagnostic process. As a result, mNGS may demonstrate better clinical performance and lead to new testing algorithms.

This proof-of-concept study was designed to establish the suitability of the UG 100 sequencing platform for metagenomic pathogen detection from clinical CSF samples. Performance characteristics included accuracy versus orthogonal results, relative pathogen nucleic acid recovery, and limit of detection.

## Materials and Methods

### CSF Sample Preparation

Residual CSF specimens from routine clinical testing performed at a national reference laboratory were selected to cover a range of possible pathogens responsible for M/E. All positive samples required at least one positive test result from a laboratory developed test or FDA-cleared assay. CSF samples negative for all targets on the FilmArray Meningitis/Encephalitis Panel (BioFire, Salt Lake City, UT) were selected to serve as negative controls and matrices for spike-in studies.

500 µL of CSF specimen was bead beat on a FastPrep24 using Lysing Matrix E (MPBio, Santa Ana, CA) at 6.0 M/S for 40 sec. After centrifugation, DNA was extracted from 200 µL of the supernatant using the Perkin Elmer Chemagen MSM1 nucleic acid extraction platform and Chemagic Viral NA/gDNA Kit (PerkinElmer, Springfield, IL) eluting in 50 µL of elution buffer.

Nucleic acid enrichment was performed using the NEBNext Microbiome DNA Enrichment Kit (New England Biolabs, Ipswich, MA) according to the manufacturer instructions. DNA extract (40 µL) was combined with 160 µL of MBD2-Fc-bound magnetic beads and 1X binding buffer. Unbound microbial DNA was put through bead cleanup using Agencourt AMPure magnetic beads (Beckman Coulter, Indianapolis, IL). Microbial DNA was eluted in 50 µL of TE.

### LOD Testing

Patient CSF specimens positive for human herpesvirus 1 (HSV-1) or *Streptococcus pneumoniae* were used for spiking and were quantified in a published real-time PCR assay for *S. pneumoniae* or an unpublished assay for HSV-1 (ELiTech, Silicon Valley, CA) from a standard curve generated using known concentrations of plasmid DNA ^14,15^. HSV-1 assay primers (HSV1-L1 (AATAAATCAGGGAGTTGTTCGGTCATAAGC) and HSV1-E2 (AATAAATCATCGGAACGCACCA*CACAAAAG)) and probe (HSV1-DSQ-AP593 (G*TAGTTGGTCGTTCGC)) were used at 0.3 µM and 0.2 µM, respectively. Super A or super G (*) bases were used to optimize primer and probe design. DNA was amplified with the QuantStudio 12k Flex Real-Time PCR System (ThermoFisher) using the following conditions: 95 °C for 10 min followed by 45 cycles of 95 °C for 10 sec, 55 °C for 45 sec. Fluorescence was read after each of the 45 cycles. Pooled CSF patient samples negative in the BioFire Meningitis/Encephalitis Panel were spiked with pathogen nucleic acid at 500,000 copies per mL and ten-fold serial dilutions to 5 copies/mL were created for assessing sensitivity.

### Ultima Genomics Sample Prep and Sequencing

DNA samples were prepared as libraries following Ultima Genomics PCR-Free WGS Library Preparation Protocol (P00014) with a varying input of 10-20ng of DNA from each sample. Due to the low genomic material of samples, preparation deviated from the protocol by diluting adapter input 1:10 prior to adapter ligation. Next libraries were PCR-amplified using Ultima Genomics Library Amplification Kit v4.0 Native User Guide (P00068) with 15 cycles of PCR instead of the recommended 7 cycles.

After library prep, the samples were sequenced on the UG 100 Sequencer using UG Baseline 1.5 Sequencing Chemistry. This chemistry generated single-end 300bp reads using the sequencing recipe UG_116cycle_Baseline_1.5.3.2. A human genome control sample HG002 (Coriell Nucleic acid ID: NA24385) was spiked into each run to ensure quality of the samples sequenced.

### Bioinformatics analyses

All non-amplifying UG primers, adapters, and barcodes were trimmed on the UG 100 server using the Ultima Genomics in house trimmer software. Reads were then aligned to the human genome assembly GRCh38/hg38 using BWA MEM^16^. Reads not aligned to the human genome were extracted from the BAM alignment and converted to fastq using samtools v1.11^17^, thereby removing all reads supposably originating from human DNA.

To assess the suitability of UG 100 sequencing reads for metagenomics, non-human reads from each sample were aligned to a reference genome of the orthogonally confirmed pathogen. Additional metagenomics was performed with Kraken2 against the pre-built NT database with default settings^18^. Antimicrobial resistance genes were examined using CARD and manually confirmed with BLAST^19^. Alignment and bioinformatics to interrogate taxonomical assignment of reads was also manually performed using Geneious Prime 2025.0.3 (https://www.geneious.com) and BLAST^20^.

## Ethics Statement

This study was approved under University of Utah Institutional Review Board (#00078952) as waived research, informed consent was not required.

## Results

### Accuracy and Sequencing Metrics

UG 100 sequencing produced reads aligning to the known pathogen in 93% (26/28) of positive cases, which included 19 unique organisms (Table 1). Of the two samples which failed detection, one (CSFU1-08) was due to a sequencing failure (no useable reads generated). In the other sample (CSFU2-13) no reads corresponding to the known pathogen (HHV-6) were identified. On a per sample basis, normalized pathogen reads per million (RPM) ranged from 2.83-15,746. Overall genomic coverage of pathogen genomes ranged from 0.1% to 98.1%, and average sequencing depth ranged from 0-304 reads.

**Table 1:** Detection of Known Pathogens in Clinical Samples.

| ID | Known Organism | Positive Orthogonal Test | Ultima Result | RPM | Coverage | Depth (Mean) | Reference Genome | Non-human reads | Aligned Reads |
| --- | --- | --- | --- | --- | --- | --- | --- | --- | --- |
| CSFU2-04 | *Streptococcus agalactiae* | Biofire panel positive | Detected | 18 | 5.40% | 0.3 | GCF_0015520 | 352,793 | 3,611 |
| CSFU2-06 | *Toxoplasma gondii* | LDT NAAT (CT 23.7) | Detected | 1,819.34 | 30.00% | 1.1 | GCF_0000065 | 331,030 | 273,753 |
| CSFU2-07 | *Toxoplasma gondii* | LDT NAAT (CT 29.1) | Detected | 818.17 | 10.50% | 0.6 | GCF_0000065 | 297,694 | 162,377 |
| CSFU2-08 | *Taenia solium* | IgG ELISA (35 IU [>11 IU is positive]) | Detected | 6,898.19 | 5.20% | 4.7 | GCA_0018707 | 2,220,401 | 1,472,764 |
| CSFU2-10 | *Cryptococcus neoformans* | Biofire panel positive | Detected | 1,385.00 | 2.20% | 3.9 | GCF_0000910 | 3,709,025 | 244,619 |
| CSFU2-13 | Human herpesvirus 6 | Biofire panel positive | Not Detected | 0 | 0.00% | 0 | NC_001664 | 6,299,977 | 0 |
| CSFU2-14 | Herpes simplex virus type 1 | Biofire panel positive | Detected | 44.89 | 63.50% | 13.9 | NC_001806.2 | 383,631 | 8,610 |
| CSFU2-15 | Herpes simplex virus type 2 | Biofire panel positive | Detected | 35.21 | 78.40% | 8.6 | NC_001798 | 299,159 | 6,196 |
| CSFU2-16 | Varicella zoster virus (HHV3) | Biofire panel positive | Detected | 7.16 | 38.90% | 2.1 | NC_001348 | 124,193 | 1,271 |
| CSFU2-17 | *Candida* species | LDT NAAT (CT 32.0) | Detected | 76.12 | 0.60% | 0.1 | GCF_0001829 | 944,670 | 14,381 |
| CSFU2-18 | *Coccidioides* species | Complement fixation 1:64, Immunodiffusion Positive | Detected | 139.84 | 0.60% | 0 | GCF_0001493 | 645,354 | 27,347 |
| CSFU2-19 | *Coccidioides* species | Complement fixation 1:32, Immunodiffusion Negative | Detected | 14.33 | 0.10% | 0 | GCF_0001493 | 5,286,913 | 4,952 |
| CSFU2-31 | *Neisseria meningitidis* | Biofire panel positive | Detected | 15,746.29 | 92.80% | 304 | NZ_CP064367 | 2,778,867 | 2,290,934 |
| CSFU2-32 | *Escherichia coli* | Biofire panel positive | Detected | 132.88 | 49.10% | 1.2 | NC_000913.3 | 89,277 | 23,726 |
| CSFU2-33 | *Haemophilus influenza* | Biofire panel positive | Detected | 20.04 | 17.50% | 0.3 | NZ_CP085952 | 67,412 | 3,377 |
| CSFU2-34 | Cytomegalovirus (HHV5) | Biofire panel positive | Detected | 2.83 | 2.20% | 0.1 | NC_006273.2 | 47,154 | 445 |
| CSFU2-35 | *Listeria monocytogenes* | Biofire panel positive | Detected | 28.99 | 1.50% | 0.2 | NC_003210.1 | 264,753 | 5,904 |
| CSFU2-36 | *Cryptococcus neoformans* | Biofire panel positive | Detected | 188.37 | 1.40% | 0.3 | GCF_0000910 | 833,540 | 38,297 |
| CSFU2-37 | *Borrelia burgdorferi* | LDT NAAT (CT 43.0) | Detected | 9.03 | 1.60% | 0.1 | NC_012512.1 | 229,929 | 2076 |
| CSFU1-02 | *Haemophilus influenzae* | Biofire panel positive | Detected | 359.80 | 97.60% | 9.3 | CP007470.1 | 121,976 | 62,726 |
| CSFU1-04 | Human herpesvirus 6 | Biofire panel positive | Detected | 3.57 | 20.40% | 1.4 | NC_001664 | 86,063 | 755 |
| CSFU1-16 | Herpes simplex virus type 1 | Biofire panel positive | Detected | 23.76 | 98.10% | 6.7 | NC_001806.2 | 94,348 | 4,684 |
| CSFU1-14 | *Streptococcus pneumoniae* | Biofire panel positive | Detected | 15.56 | 20.20% | 0.3 | CP053210 | 78,369 | 3,038 |
| CSFU1-13 | *Borrelia burgdorferi* | LDT NAAT (CT 39.7) | Detected | 6.10 | 5.40% | 0.1 | NC_012512.1 | 147,751 | 1,188 |
| CSFU1-17 | *Toxoplasma gondii* | LDT NAAT (CT 31.9) | Detected | 286.37 | 7.20% | 0.2 | NC_031467.1 | 443,146 | 54,429 |
| CSFU1-10 | *Coccidioides immitis* | Complement fixation 1:32 | Detected | 9.80 | 1.10% | 0 | NW_0045043 | 96,906 | 1,721 |
| CSFU1-12 | *Taenia solium* | IgG ELISA (24 IU [>11 IU is positive]) | Detected | 145.54 | 1.00% | 0 | GCA_0020824 | 180,312 | 23,329 |
| CSFU1-08 | Cytomegalovirus (HHV5) | LDT NAAT (CT 29.4) | Not Detected | 0 | 0 | 0 | NC_006273.2 | 0 | 0 |

For clinical samples, the average number of total reads per sample was 190 million (range: 145M – 345M) (Table S1). Across all clinical samples, the average number of non-human reads was 979,802 (47,154 – 6,299,977) with an average quality score or 32.2 (31.7-32.7). The average read length was 258 nucleotides (mean per-sample read length 221-281).

### Specificity

For the sixteen clinical samples tested on the BioFire M/E panel, specificity analysis was performed. Non-human reads were aligned against reference genomes of the organisms included on the panel. When an analyte reported as “not detected” on the M/E panel was observed at >3 RPM by mNGS, results were classified as discordant. In 75% (12/16) of specimens, at least one target analyte reported as “not detected” by the BioFire M/E panel had >3 RPM detected by mNGS (Table S2). The most frequently identified was *E. coli* (12/16) and *S. pneumoniae* (6/16).

### Limit of Detection Study

Using serially diluted HSV-1 samples, 77 non-identical reads (0.48 RPM) mapping to the HSV-1 genome were recovered at 50 copies/mL, below the 100 copies/mL LoD for the commercially available NAAT. For *S. pneumoniae*, 6,777 reads (35.69 RPM) mapping to the reference genome were recovered at 5 copies/mL which is below the published LoD of 1,500 copies/mL for the NAAT^14^ (Figure 1).

**Figure 1:**
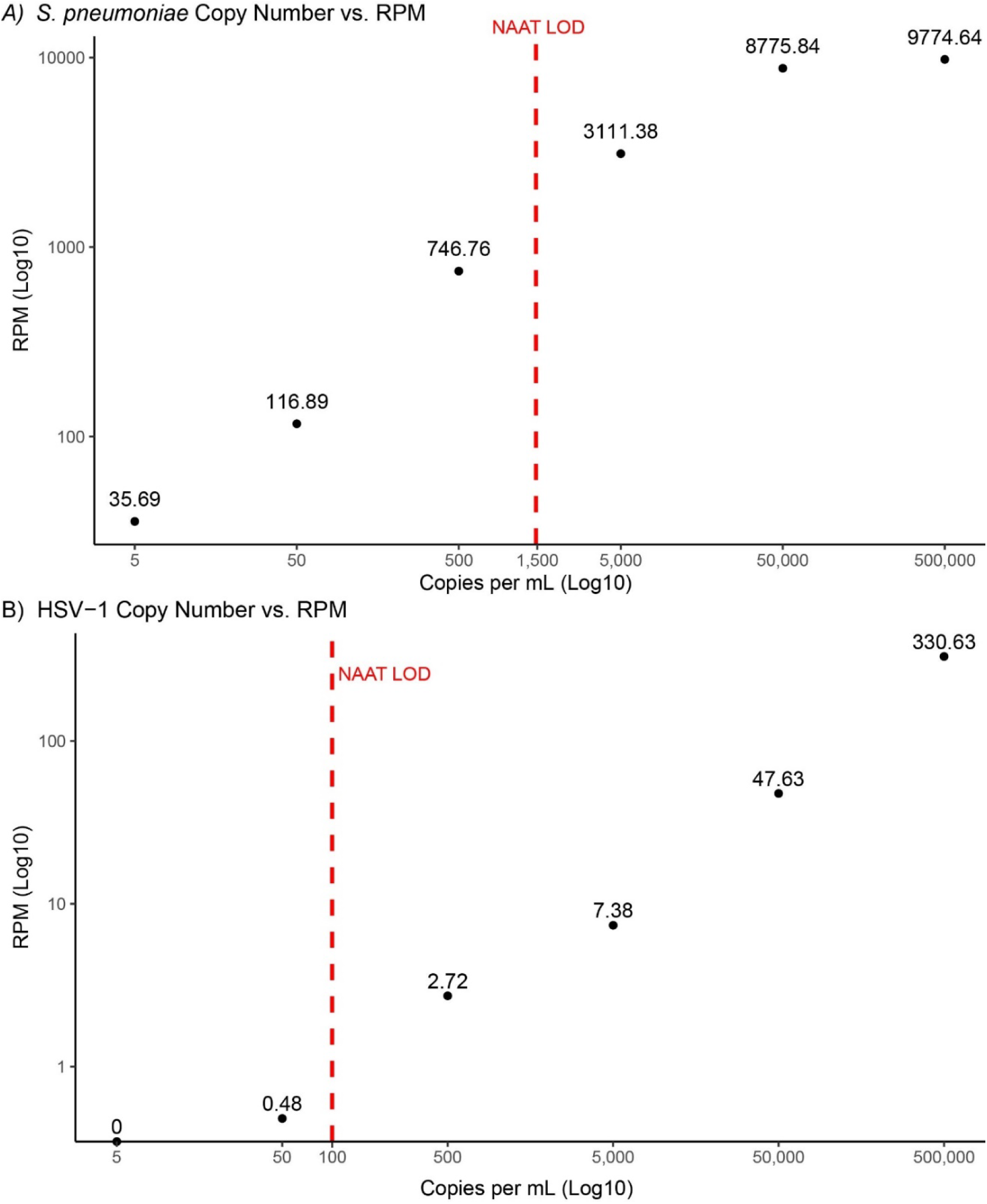
Limit of Detection for HSV-1 and *Streptococcus pneumoniae* by mNGS on the Ultima UG 100. A) A total of 6,777 reads (35.69 RPM) aligning to the *S. pneumoniae* genome, were recovered at 5 copies/mL well below the published LoD of 1,500 copies/mL for the NAAT (red dotted line). B) A total of 77 reads (0.48 RPM) mapping to the HSV-1 genome were recovered at 50 copies/mL, below the 100 copies/mL LoD for the commercially available NAAT (red dotted line).

### Novel Observations

Near complete genomes were recovered from three positive samples; CSFU1-02 (*Haemophilus influenza*; 359 RPM, 97.6% genome coverage, average depth 9.3 reads) (Figure 2A), CSFU1-16 (HSV-1; 23.76 RPM, 98.1% genome coverage, average depth 6.7 reads), and CSFU2-31 (*Neisseria meningitidis*; 15,746 RPM, 92.80% genome coverage, average depth 304 reads). Further analysis of the *H. influenza* sequence identified putative antimicrobial resistance genes, *hmrM* and *lpsA*.

**Figure 2:**
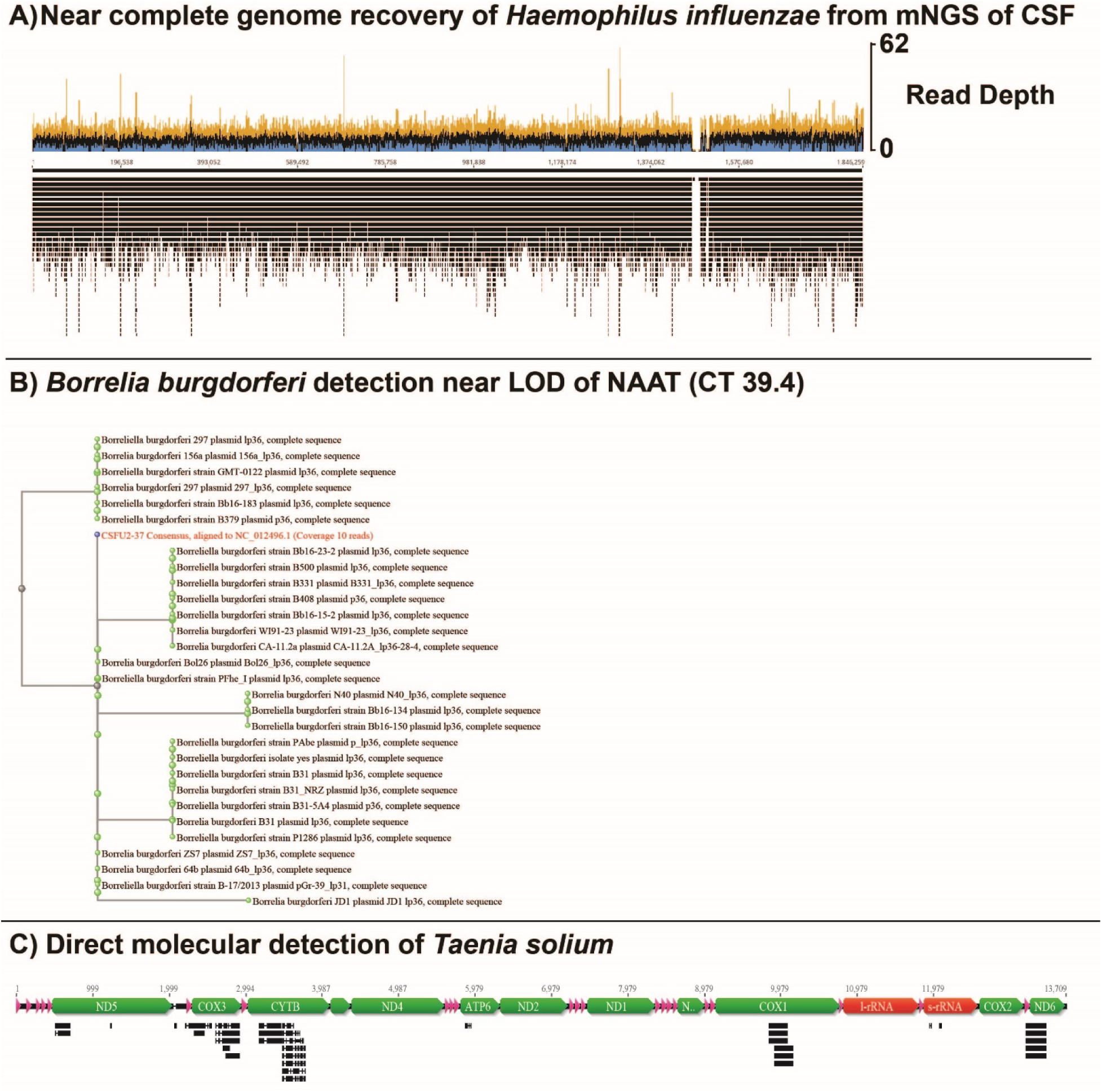
Novel Observations Following mNGS with Ultima Sequencing Chemistry. <u>A) Near-complete genome recovery of *Haemophilus influenzae* from mNGS of CSF</u> – Recovery of a near-complete genome of *Haemophilus influenzae* (97.6% genome coverage, average depth 9.3 reads) directly from CSF. <u>B) *Borrelia burgdorferi* detection near LOD of NAAT (CT 39.4)</u> – Phylogenetic tree of *Borrelia burgdorferi* using reads recovered (2076 reads, 6.1 RPM) from a clinical sample near the limit of detection (CT 39.4). <u>C) Direct molecular detection of *Taenia solium*</u> – Reads (n=13,409) mapping to specific mitochondrial sequences of *Taenia solium* recovered from patient with positive serology results.

Two samples containing low levels of *Borrelia burgdorferi* DNA (CT value of 39.7 for CSFU1-13 and 43.0 for CSFU2-37) as determined by a laboratory-developed test were positively identified by mNGS at an RPM value of 6.10 and 9.03, respectively (Figure 2B). Ultima sequencing of two samples (CSFU1-12 and CSFU2-08) with positive serologic evidence for *Taenia solium* infection (IgG ELISA of 24 IU and 35 IU) detected reads corresponding to the *T. solium* genome in both cases, at 145.54 RPM and 6,898.19 RPM, respectively. As the publicly available genomes for *T. solium* are poorly annotated, reads corresponding specifically to the *T. solium* mitochondrial sequences were manually confirmed using BLAST (Figure 2C).

## Discussion

This proof-of-concept study was designed to assess the suitability of the UG 100 sequencing platform for metagenomic pathogen detection in cases of M/E. Overall, we found suitable performance characteristics as compared to existing assays with a sensitivity of 93%. In one case (CSFU1-08, CMV), DNA extraction and amplification failed, preventing generation of sequencing reads and analysis. The other case in which Ultima sequencing failed to detect a pathogen (CSFU2-13, HHV-6), zero reads were recovered that mapped to the HHV-6 reference genome. Basic metagenomic analysis of this sample with Kraken2 revealed many reads (8,454) aligning to HSV-1 in this sample. This sample’s reads were manually aligned to the HSV-1 reference genome (NC_001806) and 7,586 reads were identified. However, these reads were restricted to only a few sites on the genome, suggesting an analytic error (i.e. jackpotting) and showed 100% nucleotide agreement to the HSV-1 isolate used for LOD studies on the same run. Additionally, this sample contained 41,958 reads mapping to *Cryptococcus neoformans.* CSFU2-13 was located next to CSFU2-10 which contained a relatively high amount of pathogen DNA (244,619 reads mapping to *C. neoformans).* Unfortunately, the initial DNA extraction for mNGS depleted the sample, and further analysis with re-sequencing was not possible. As this sample had a positive NAAT for HHV-6 and we recovered zero reads aligning to HHV-6, we suspect a pre-analytic error such as a sample swap with a negative, combined with contamination events exacerbated by physical plate proximity to samples with high pathogen DNA content.

Specificity analysis was performed in a subset of samples for which BioFire M/E panel results were available. Analytes present on the panel but reported as “Not Detected” were used for comparison to mNGS results. Off-target reads were detected in every sample; however, using a >3 RPM cutoff 75% (12/16) of samples contained a discordant organism. The most common discordant detection was *E. coli* (12/16). As specificity was analyzed by aligning the sample’s non-human reads to the reference genome of each M/E panel target, these reads likely represent shared bacterial sequence from environmental, skin, container, and reagent components^21^. Mitigating the effects of these DNA sources remains a challenge metagenomic analysis. To illustrate this point, *Cutibacterium acnes* nucleic acid was detected at >3RPM in 94% (15/16) of samples. After *E. coli*, the next two most common M/E panel organisms detected discordantly were *S. pneumoniae* in 38% (6/16) of samples and *H. influenza* in 25% (4/16) of samples. Both of these organisms were present in other clinical samples on the runs at very high concentrations. These findings highlight the need for both in-run and between-run background normalization, as the extremely sensitive nature of metagenomics exacerbates any potential contamination events.

This study also provided several new observations in how a low-cost, high-sequencing depth mNGS assay may improve upon existing options. First, we recovered near-complete pathogen genomes from three clinical samples. Significantly increasing the number of reads per sample when performing mNGS opens new avenues of analysis including identification of antimicrobial resistance genes and strain typing for outbreak tracing. We also showed molecular detection of an organism (*Taenia solium*) for which no FDA-cleared molecular assays are available. By reducing the sequencing expense of clinical mNGS, pathogens which have historically required esoteric molecular testing through public health labs or serologic assays for diagnosis may be identified earlier and more precisely with mNGS. Lastly, we demonstrate that our mNGS workflow provides a limit of detection that may surpass that of single target NAATs. This highly sensitive detection of pathogen DNA fragments was evidenced in both contrived samples and two clinical samples positive for *B. burgdorferi* with late cycle threshold values.

## Flipping the Diagnostic Algorithm and Cost Comparisons

Historically, mNGS is most likely to be ordered when the traditional diagnostic armamentarium has been exhausted by negative results. Interestingly, many traditional microbiology tests ordered early in the diagnostic workflow also demonstrate low clinical sensitivity but are still used because they are inexpensive and readily available. For example, a single blood culture has between 70-80% sensitivity and costs between $1-10^22^. If the cost of mNGS assays were dramatically reduced, a potential “flipping” of the diagnostic workflow becomes possible. In this new workflow, mNGS is ordered earlier or as an up-front “rule out” test for infectious diseases.

Cost comparison for this sequencing study compared favorably to the current industry standard. The approximate cost per 50M reads would be $12 using Ultima chemistry which is 25% of the cost of Illumina NovaSeqX sequencing ($48 per 50M reads). Additionally, for the purposes of diagnosing of M/E by mNGS, the cost of DNA extraction and library prep would be expected to be essentially identical between the two platforms. Lowered sequencing costs may reduce the need to batch samples which could lead to shorter turn-around-times. In this study we targeted $60 in total sequencing costs (or 250M reads per sample) and another $40 in reagent costs to bring the total cost to perform mNGS to $100.

It is important to note that following basic QC and trimming, FASTQ files from the UG 100 sequencing platform can be used with a range of existing bioinformatics tools (FastQC, Geneious, bowtie, MAAFT, Blast, and Kraken2). The ability to “plug and play” with reads from a novel sequencing technology is highly useful as it significantly reduces the bioinformatics and analytic overhead of adopting this novel technology.

## Study Limitations and Conclusion

Limitations of this study include manual selection of clinical samples, restriction to DNA-based organisms, lack of prospective design, and a lack of robust metagenomic analysis. As in any metagenomic study, reads were recovered to a wide variety of off target organisms. However, development and validation of a novel metagenomic analysis pipeline was determined to be beyond the scope of this study. Therefore, manual bioinformatics and basic use of pre-existing tools were used to determine if reads from this sequencer would be useable by other, more sophisticated metagenomic pipelines.

The UG 100 sequencing platform offers a new NGS chemistry allowing for low cost metagenomic pathogen identification. Overall accuracy was 93% with a LOD that surpassed single-target NAATs. While further bioinformatic refinements are necessary before entering the clinical diagnostic space, this proof-of-concept study illustrates the technical reality needed for first-line mNGS testing of patients is approaching.

## Data Availability

All data produced in the present study are available upon reasonable request to the authors

## Data Availability

All data produced in the present study are available upon reasonable request to the authors

---

**Supplemental Table 1:** Run Characteristics of Clinical Samples.

| Supplemental Table 1: Run Characteristics of Clinical Samples |  |  |  |  |  |  |
| --- | --- | --- | --- | --- | --- | --- |
| ID | Total Reads | Human (%) | Human Reads | Non-human read | Mean Quality | Mean Read Length |
| CSFU2-04 | 195,995,871 | 99.82 | 195,643,078 | 352,793 | 31.83 | 271.06 |
| CSFU2-06 | 150,468,095 | 99.78 | 150,137,065 | 331,030 | 31.9 | 264.48 |
| CSFU2-07 | 198,462,837 | 99.85 | 198,165,143 | 297,694 | 32.58 | 221.72 |
| CSFU2-08 | 213,500,059 | 98.96 | 211,279,658 | 2,220,401 | 32.67 | 222.25 |
| CSFU2-10 | 176,620,247 | 97.9 | 172,911,222 | 3,709,025 | 32.08 | 256.22 |
| CSFU2-13 | 155,554,999 | 95.95 | 149,255,022 | 6,299,977 | 32.14 | 256.08 |
| CSFU2-14 | 191,815,311 | 99.8 | 191,431,680 | 383,631 | 32.14 | 256.05 |
| CSFU2-15 | 175,976,074 | 99.83 | 175,676,915 | 299,159 | 32.43 | 243.96 |
| CSFU2-16 | 177,419,232 | 99.93 | 177,295,039 | 124,193 | 32.46 | 243.29 |
| CSFU2-17 | 188,933,930 | 99.5 | 187,989,260 | 944,670 | 32.14 | 257.65 |
| CSFU2-18 | 195,561,952 | 99.67 | 194,916,598 | 645,354 | 32.04 | 266.51 |
| CSFU2-19 | 345,549,853 | 98.47 | 340,262,940 | 5,286,913 | 32.43 | 242.84 |
| CSFU2-31 | 145,490,413 | 98.09 | 142,711,546 | 2,778,867 | 31.74 | 280.82 |
| CSFU2-32 | 178,554,716 | 99.95 | 178,465,439 | 89,277 | 31.93 | 269.56 |
| CSFU2-33 | 168,529,684 | 99.96 | 168,462,272 | 67,412 | 31.93 | 270.12 |
| CSFU2-34 | 157,179,466 | 99.97 | 157,132,312 | 47,154 | 32.24 | 252.9 |
| CSFU2-35 | 203,656,477 | 99.87 | 203,391,724 | 264,753 | 32.02 | 258.45 |
| CSFU2-36 | 203,302,407 | 99.59 | 202,468,867 | 833,540 | 32.04 | 266.95 |
| CSFU2-37 | 229,929,499 | 99.9 | 229,699,570 | 229,929 | 31.88 | 270.54 |
| CSFU1-02 | 174,334,763 | 99.93 | 174,212,787 | 121976 | 32.24 | 279.53 |
| CSFU1-04 | 211,429,994 | 99.96 | 211,343,931 | 86063 | 31.81 | 270.29 |
| CSFU1-16 | 197,119,035 | 99.95 | 197,024,687 | 94348 | 32.34 | 260.74 |
| CSFU1-14 | 195,189,193 | 99.96 | 195,110,824 | 78369 | 32.29 | 267.75 |
| CSFU1-13 | 194,635,540 | 99.92 | 194,487,789 | 147751 | 32.25 | 267.98 |
| CSFU1-17 | 190,066,351 | 99.77 | 189,623,205 | 443146 | 32.5 | 248.12 |
| CSFU1-10 | 175,675,005 | 99.94 | 175,578,099 | 96906 | 32.33 | 263.68 |
| CSFU1-12 | 160,288,267 | 99.89 | 160,107,955 | 180312 | 32.33 | 262.03 |
| CSFU1-08 | - | - | - | - | - | - |

**Supplemental Table 2:** Samples with known M/E panel positives aligned to all targets on panel (Reads per Million)

|  | CSFU1-02 | CSFU1-04 | CSFU1-14 | CSFU1-16 | CSFU2-04 | CSFU2-10 | CSFU2-13 | CSFU2-14 |
| --- | --- | --- | --- | --- | --- | --- | --- | --- |
|  | *Haemophilus influenza* | *Human herpesvirus 6* | *Streptococcus pneumoniae* | *Herpes simplex virus type 1* | *Streptococcus agalactiae* | *Cryptococcus neoformans* | *Human herpesvirus 6* | *Herpes simplex virus type 1* |
| *Cryptococcus neoformans* | 0 | 0 | 0 | 0 | 1 | 1,385 | 22 | 0 |
| *Escherichia coli* | 16 | 0 | 12 | 11 | 9 | 168 | 404 | 11 |
| Enterovirus | - | - | - | - | - | - | - | - |
| *Haemophilus influenza* | 360 | 1 | 1 | 1 | 2 | 20 | 27 | 3 |
| Herpes simplex virus type 1 | 1 | 0 | 1 | 24 | 0 | 0 | 54 | 45 |
| Herpes simplex virus type 2 | 1 | 0 | 1 | 10 | 0 | 1 | 28 | 25 |
| Varicella zoster virus (HHV3) | - | 0 | - | 0 | 0 | 0 | - | 0 |
| Cytomegalovirus (HHV5) | 0 | 0 | 0 | 0 | 0 | 3 | 1 | 0 |
| *Human herpesvirus 6* | 1 | 4 | 1 | 0 | 2 | 2 | - | 1 |
| *Listeria monocytogenes* | 1 | 0 | 0 | 0 | 1 | 9 | 21 | 1 |
| *Neisseria meningitidis* | 6 | 1 | 0 | 0 | 2 | 39 | 58 | 4 |
| *Human parechovirus* | - | - | - | - | - | - | - | - |
| *Streptococcus agalactiae* | 1 | 1 | 1 | 0 | 18 | 17 | 15 | 2 |
| *Streptococcus pneumoniae* | 1 | 2 | 16 | 0 | 4 | 45 | 60 | 4 |
|  | CSFU2-15 | CSFU2-16 | CSFU2-31 | CSFU2-32 | CSFU2-33 | CSFU2-34 | CSFU2-35 | CSFU2-36 |
|  | *Herpes simplex virus type 2* | *Varicella zoster virus (HHV3)* | *Neisseria meningitidis* | *Escherichia coli* | *Haemophilus influenza* | *Cytomegalovirus (HHV5)* | *Listeria monocytogenes* | *Cryptococcus neoformans* |
| *Cryptococcus neoformans* | 1 | 1 | 1 | 1 | 1 | 1 | 1 | 188 |
| *Escherichia coli* | 11 | 4 | 131 | 133 | 2 | 1 | 7 | 16 |
| Enterovirus | - | - | - | - | - | - | - | - |
| *Haemophilus influenza* | 1 | 1 | 191 | 2 | 20 | 0 | 2 | 2 |
| Herpes simplex virus type 1 | 23 | 0 | 0 | 0 | 0 | 0 | 0 | 0 |
| Herpes simplex virus type 2 | 35 | 0 | 0 | 0 | 0 | 0 | 0 | 0 |
| Varicella zoster virus (HHV3) | 0 | 7 | 0 | 0 | 0 | 0 | - | 0 |
| Cytomegalovirus (HHV5) | 0 | 0 | 0 | 0 | 0 | 3 | 0 | 0 |
| *Human herpesvirus 6* | 1 | 1 | 1 | 0 | 1 | 0 | 1 | 1 |
| *Listeria monocytogenes* | 1 | 1 | 33 | 0 | 0 | 0 | 29 | 3 |
| *Neisseria meningitidis* | 2 | 1 | 15,746 | 1 | 0 | 0 | 2 | 2 |
| *Human parechovirus* | - | - | - | - | - | - | - | - |
| *Streptococcus agalactiae* | 1 | 1 | 36 | 0 | 0 | 0 | 0 | 3 |
| *Streptococcus pneumoniae* | 2 | 1 | 39 | 0 | 0 | 0 | 2 | 8 |
| Over 3RPM off target |  |  |  |  |  |  |  |  |
| Expected Positive |  |  |  |  |  |  |  |  |

